# *TET2*-mutant clonal hematopoiesis alters neutrophil maturation and granularity

**DOI:** 10.64898/2026.11.17.26368301

**Authors:** Elina K. Cook, Marco M. Buttigieg, Yi Z. Luo, Andrew Tian, Akshaj Darbar, Caitlyn Vlasschaert, Richard N. Armstrong, Amy J. M. McNaughton, David Good, Rena Buckstein, Graeme R. Quest, Jacqueline Boultwood, Alexander G. Bick, Michael J. Rauh

## Abstract

The effects of clonal hematopoiesis (CH) on neutrophil development and function in human blood remain largely unexplored. Here, we use primary peripheral blood and bone marrow samples to characterize changes in neutrophil maturation and granule production that occur in CH.

## Background

Neutrophils are the most abundant leukocytes in the human body and play a central role in orchestrating the innate response to microbial infections, whose dysfunction is implicated in chronic inflammation and age-related pathophysiology.^1,2^ The production of neutrophils is sustained by the differentiation of hematopoietic stem and progenitor cells (HSPCs) in the bone marrow and spans various developmental stages.^1,2^ HSPCs that develop into myeloblasts commit to the granulocyte lineage, progressing through the promyelocyte, myelocyte, and metamyelocyte stages prior to becoming mature neutrophils.^1,2^ These differentiating granulocytes undergo the tightly regulated process of granulopoiesis, in which cells sequentially acquire the azurophilic (primary), specific (secondary), and gelatinase (tertiary) granules, and secretory vesicles that mediate critical neutrophil effector functions.^1,2^

Defective granulopoiesis is a long recognized feature of leukemia,^3,4^ and the resulting inflammation and loss of anti-microbial function synergize with chemotherapy-induced neutropenia to drive infectious comorbidity and mortality in patients with acute leukemia and myelodysplasia.^5–7^ Clonal hematopoiesis (CH) is the pre-leukemic acquisition of somatic mutations in HSPCs, leading to disproportionate production of mature immune progeny which inherit the clonal mutation.^8,9^ CH mutations most commonly impact epigenetic regulators like *DNMT3A* and *TET2*, which can have functional consequences leading to sustained inflammation, tissue damage, and chronic disease.^10,11^ The inflammatory pathobiology of CH has largely been studied in the context of monocyte/macrophages; however, a limited set of recent studies have appreciated the effects of CH mutations on the differentiation, migration, and effector function of neutrophils.^12–16^ Here, we systematically profile human neutrophils in CH across the bone marrow and peripheral blood, identifying *TET2*-associated alterations in neutrophil maturation and granularity with relevance to human infectious disease.

## Results

Profiling of whole blood from age-, sex-, and comorbidity-balanced patients recruited at Sunnybrook and Baycrest Hospitals (n=30; Cohort 1; Supplemental Table 1) with bulk RNA-sequencing revealed significant upregulation of neutrophil and granule related genes and pathways amongst CH+ participants (e.g. *DEF4A, CRISP3, BPI, LTF*; Supplemental Figure 1; Supplemental Table 2). These expression changes were driven almost entirely by *TET2*-mutant CH, whereas participants with *DNMT3A*-mutant CH saw no significant difference from CH-free controls (Figures 1A-B; Supplemental Figure 2; Supplemental Tables 3-4). Complete blood count data demonstrated no difference in absolute neutrophil count by CH status or any CH driver gene (Supplemental Figure 3), as demonstrated in prior works.^8,17,18^ Expression of granule component genes is sequentially regulated during granulopoiesis to create the distinct granule classes found in mature neutrophils (i.e. targeting by timing), and thus the mRNA expression of neutrophil granule components may not necessarily reflect their protein abundance.^19^ We therefore evaluated the elevated serum levels of granule proteins, lactotransferrin (LTF) and myeloperoxidase (MPO), in CH with ELISA. Despite the transcriptional effects of CH being largely driven by *TET2*, serum LTF and MPO levels were broadly upregulated across CH driver genes (Figures 1C-D), though serum and intracellular granule protein levels may differ. The distinction between mRNA and protein expression of granule components in CH may also suggest a more low-density/immature-like phenotype in circulating neutrophils.^19–21^ We thus queried differentially expressed genes from *TET2*-mutant CH in NeuMap,^22^ a transcriptional map of neutrophil development, identifying a significant enrichment of immature neutrophil markers and additional overlap with pre-neutrophil genes (Supplemental Table 5). Transcription factor activity estimation similarly showed C/EBP-ε – a key regulator of the promyelocyte-to-myelocyte transition^1^ – among the top positively enriched transcription factors in *TET2*-mutant CH (Supplemental Figure 4). Taken together, our data suggest that *TET2* mutations shift peripheral blood neutrophils towards a more immature phenotype that is enriched for expression of certain early granule components including azurophilic and specific granule genes.

**Figure 1:**
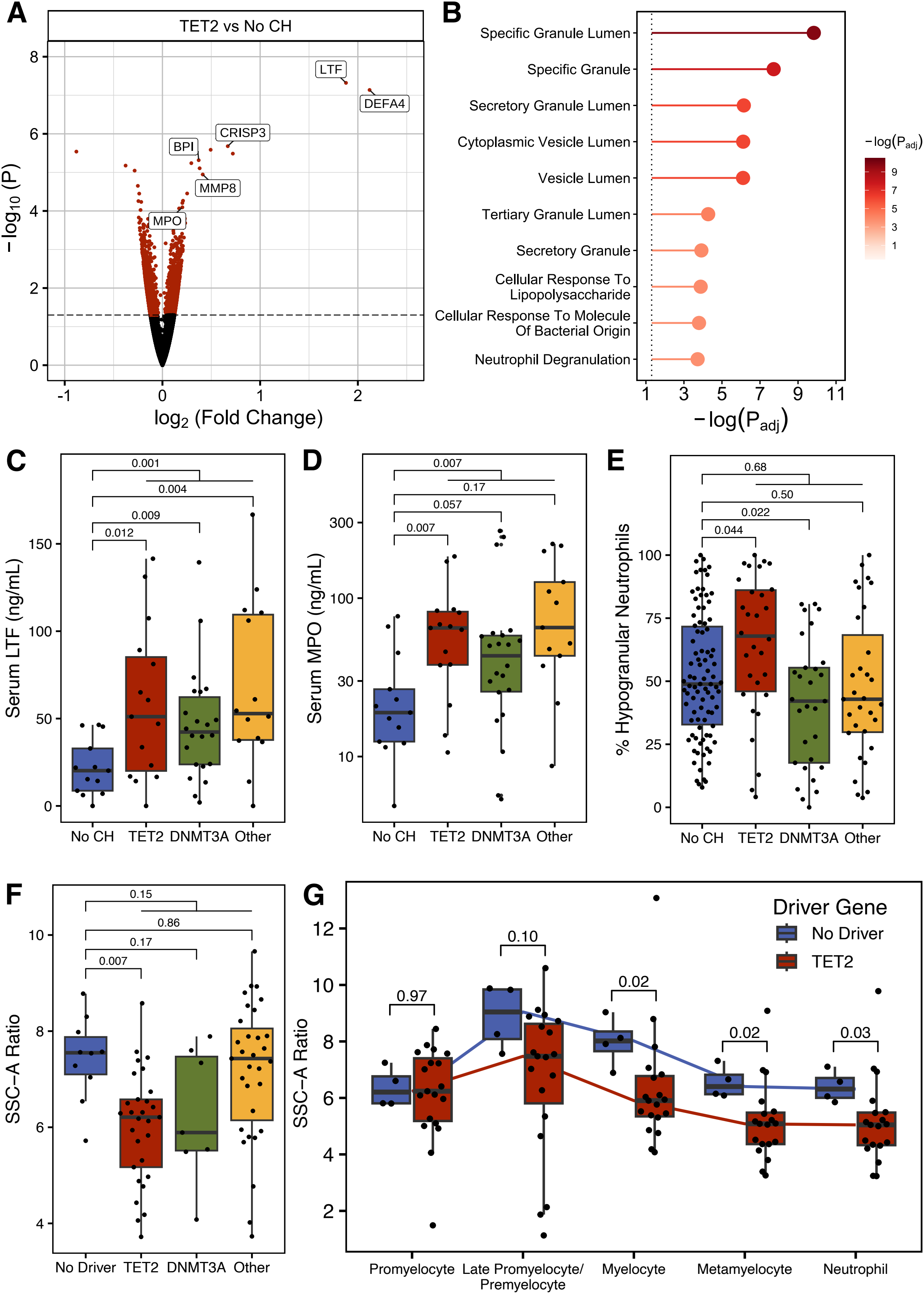
*TET2*-mutant CH is associated with altered neutrophil maturation and granule production. A) Differential gene expression in whole blood from *TET2*-mutant CH and CH-free participants. Select neutrophil and granule related genes are labelled, and coloured points represent nominal P value < 0.05. B) Gene Ontology (GO) term enrichment of the top 10 significantly enriched gene sets in *TET2*-mutant CH versus no CH. C) ELISA quantification of serum LTF levels. P values from Wilcoxon rank sum test. D) ELISA quantification of serum MPO levels. Y-axis is log_10_-scaled for visualization. P values from Wilcoxon rank sum test. E) Fraction of neutrophils identified as relatively hypogranular from peripheral blood slide images by the support vector machine model. P values from beta-binomial regression models adjusted for age, sex, and comorbidities. F) Side Scatter-A (SSC-A) ratio of granulocytes to lymphocytes from bone marrow flow cytometry. P values from linear regression models adjusted for age and sex. G) Side scatter-A (SSC-A) ratio of various granulocyte maturation states in bone marrow samples with P/LP *TET2* variants and no identifiable driver. P values from Wilcoxon rank sum test.

To expand our characterization of neutrophil maturation and granularity in CH, we then profiled peripheral blood slide images and bone marrow flow cytometry data routinely collected from patients managed at the Kingston Health Sciences Centre (KHSC). First, we developed a support vector machine learning model to quantify the relative granularity of neutrophils in peripheral blood slide images (Supplemental Figures 5-7; Methods). We then deployed this model on peripheral blood slide images from 180 patients for whom we also ascertained CH status (Cohort 2; Supplemental Table 6), to quantify the proportion of relatively hypogranular neutrophils in each sample. After adjusting for age, sex, and comorbidities, we found that *TET2*-mutant CH was associated with greater proportion of relatively hypogranular neutrophils compared to no CH, while other CH mutations nominally trended towards having fewer relatively hypogranular neutrophils (Figure 1E). Furthermore, in 77 bone marrow samples profiled with flow cytometry during clinical investigation of unexplained cytopenias or myeloid neoplasms (Cohort 3; Supplemental Table 7), granulocytes from individuals with somatic *TET2* mutations showed a significant reduction in cytoplasmic granularity by side scatter-A (SSC-A) ratio compared to individuals with no identified driver mutation (Figure 1F). SSC-A ratio also displayed a negative association with total *TET2* VAF dosage in these patients (Supplemental Figure 8). To determine the maturation state at which the granularity of *TET2*-mutant cells diverges from the reference, we analyzed data from a subset of 23 individuals who received additional diagnostic flow cytometry for markers of granulocyte maturation. Cells from *TET2*-mutant bone marrow began to nominally show a reduction in granularity by SSC-A in late promyelocytes/premyelocytes, with sustained significant reduction in granularity in myelocytes and metamyelocytes through to mature neutrophils (Figure 1G).

Finally, we assessed the potential clinical relevance of CH-mediated neutrophil granule component changes in the UK Biobank cohort, following our previous report of an association between CH and infection with bacterial pneumonia.^14^ To determine the extent that neutrophil granules may mediate this susceptibility, we leveraged the rs2333227 (G/C-463A/T) polymorphism in *MPO*, a critical component of azurophilic granules that was enriched in CH+ samples from our dataset. rs2333227-T is a common polymorphism in the *MPO* promoter that disrupts SP1 transcription factor binding and reduces *MPO* expression in neutrophils and macrophages.^23^ We found that rs2333227-T alleles specifically conferred protection from infection-related mortality associated with CH, while having no impact on the association of CH with all-cause mortality (Figure 2).

**Figure 2:**
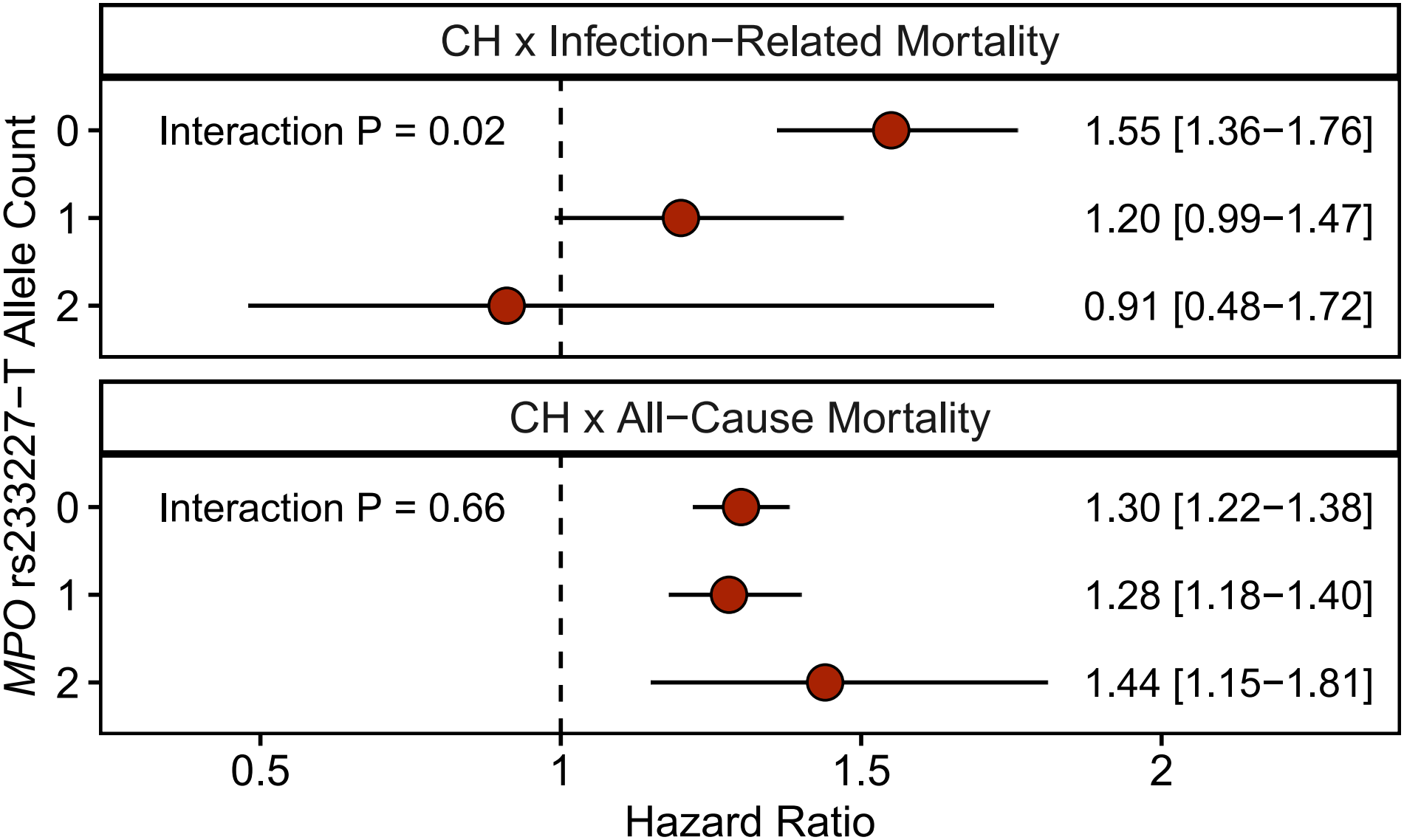
*MPO* rs2333227-T genotype mediates the association between CH and infection-related mortality. Cox proportional hazard ratios and 95% confidence intervals for CH with infection-related and all-cause mortality based on *MPO* rs2333227-T genotype in the UK Biobank.

## Discussion

Our study provides multifaceted evidence from clinical blood and bone marrow samples that altered neutrophil maturation and granule production are characteristic of human CH, especially with *TET2* mutations. We observe that CH-mutant neutrophils display a relative hypogranular phenotype in the blood and bone marrow, while reflecting greater immaturity characterized by more azurophilic and specific granule components, including myeloperoxidase and lactotransferrin. While we posit that the reduced granularity of mature neutrophils in CH is secondary to defective neutrophil maturation and granulopoiesis, it is also possible that CH-mutant neutrophils more hastily undergo degranulation in response to infectious and inflammatory stimuli. Previous studies of *TET2* loss-of-function in human neutrophils by Huerga-Encabo *et al*.^13^ have similarly identified an immature, low-density neutrophil phenotype enriched for primary granule components, however, reliance on humanized mouse xenografts limits the applicability of these findings. By elucidating CH-associated neutrophil granule and maturation changes and their roles in disease pathogenesis using primary human samples, our data supports the real-world translational relevance of neutrophils in CH-associated chronic illnesses.

Beyond the pathophysiological relevance of neutrophil changes in CH, our findings indicate that altered neutrophil phenotypes could also serve as a screening tool to aid in the identification of patients who would benefit from CH testing alongside other clinical tools such as the CH risk score (CHRS), PRISM, or CH-Predict.^18,24,25^ Our KHSC cohorts consisted largely of patients who underwent routine hematopathology assessments, suggesting that granularity assessment using accessible hematology analyzer and blood imaging data can help triage at-risk individuals for additional molecular testing as indicated.

In summary, we show that *TET2*-mutant CH drives an immature, azurophilic/specific granule-enriched phenotype in neutrophils from human peripheral blood and bone marrow samples. Additionally, we underscore the clinical significance of CH-associated changes in neutrophil granule components through their ability to mediate the epidemiologic association between CH and infection-related mortality. Future mechanistic studies are required to elucidate the regulatory mechanisms that link *TET2* and other CH driver genes to neutrophil development and effector function in healthy and diseased states.

## Methods

### Sunnybrook-Baycrest Hospital Cohort (Cohort 1)

#### Cohort Information

Samples for DNA-seq, RNA-seq, and serum ELISA were obtained from 359 communitydwelling, haematologically healthy adults ≥65 years of age at Baycrest Health Sciences and the Family Practice Unit at Sunnybrook Health Sciences Centres (Toronto, Canada) from 2014 to 2017 as described previously.^10^ Samples were collected in accordance with the Helsinki Declaration, and participants gave written informed consent to the study as approved by health sciences research ethics boards at Baycrest Hospital, Sunnybrook Hospital, and Queen’s University.

#### DNA Sequencing

DNA sequencing and CH variant calling are described extensively by Cook *et al*. ^10^ Briefly, DNA was extracted from the whole peripheral blood (PAXgene, Qiagen) following the manufacturer’s instructions. Libraries were produced targeting 48 commonly mutated genes in myeloid neoplasms, and 15-20g/library was sequenced on the Ion Proton (Thermo Fisher Scientific). Somatic variant calling was performed with IonTorrent software, and variants were manually verified using Integrative Genomics Viewer. The presence of CH was defined by putative driver mutations with a variant allele frequency (VAF) ≥0.02.

#### Bulk RNA Sequencing

A subset of 30 age-, sex-, and comorbidity-balanced cases (*TET2* n=11, *DNMT3A* n=8, Other CH n=2, No CH n=9; Supplemental Table 1) were selected for transcriptional profiling. Bulk RNA-seq was performed on ribo-depleted RNA extracted from whole peripheral blood samples (PAXgene, Qiagen) on the Illumina HiSeq 4000 platform. All samples achieved RIN >7 prior to sequencing. Sequencing was performed with 75-bp paired-end reads, achieving a depth of greater than 50 million reads per sample. Transcripts were pseudo-aligned and quantified with kallisto^26^ 0.51.1 and aggregated to the gene level with tximport^27^ 1.34.0 using GENCODE v49. Differential expression analysis with DESeq2^28^ 1.46.0 was used to compare CH and specific CH driver mutations to the CH-free reference group. Genes required a count of ≥10 in at least 5 samples to be included in the analysis. Adjusted p values were calculated using independent hypothesis weighting^29^ 1.34.0 requiring a p_adj_ <0.1 for statistical significance. Adaptive shrinkage using ashr^30^ 2.2-63 was applied to log fold change values to account for differences observed in high variance and/or low count outlier genes. Volcano plots of differential gene expression were generated with EnhancedVolcano 1.24.0. Functional pathway enrichment analysis was conducted using gProfiler2 gost^31^ 0.2.4 following default parameters using gene ontology terms and NeuMap^22^ markers. Transcription factor activity estimation was run with univariate linear modelling in decoupleR^32^ 2.12.0 using the CollecTRI^33^ network transcription factor target data based on differential expression test statistics.

#### Serum Protein Quantification

In an expanded and overlapping subset of 64 cases (CH n=51, No CH n=13), serum MPO and LTF levels were quantified in duplicate (Human MPO Instant ELISA Kit, Invitrogen; Human LTF ELISA Kit, Sigma) according to the manufacturer’s instructions with a VersaMax microplate spectrophotometer (Molecular Devices). Differences in serum MPO and LTF based on CH status were evaluated with the Wilcoxon rank sum test.

### Kingston Health Sciences Centre Cohorts (Cohorts 2 & 3)

#### Cohort Information

With Queen’s University/Kingston Health Sciences Centre (KHSC) Health Sciences Research Ethics Board approval, demographic data (age, sex, complete blood counts and parameters, and history of iron deficiency, kidney dysfunction, and liver dysfunction, or pre-existing myeloid neoplasm), residual EDTA CBC tube samples, and CellaVision digital blood cell images were collected from 362 patients undergoing standard-of-care treatment at KHSC (Kingston, Canada) as part of a larger research project (Cohort 2).

Data was also collected retrospectively from 77 patients who received routine bone marrow flow cytometry and genetic testing for myeloid cancer driver mutations at KHSC to evaluate unexplained cytopenia, clonal cytopenia of undetermined significance, and myeloid neoplasms enriched for myelodysplastic neoplasms between 2023 and 2024 (Cohort 3).

#### DNA Sequencing – Peripheral Blood Imaging Subcohort (Cohort 2)

EDTA-anticoagulated CBC blood specimens were frozen in a −80°C freezer within 48 hours of collection and before DNA extraction. The DNeasy Blood & Tissue Kit was used for DNA extraction from whole blood following the manufacturer’s protocol (Qiagen). Extracted DNA was sequenced for CH variants using a high-throughput, targeted NGS assay at the VANTAGE Core Genomics Facility as described previously.^34^ 90 subjects with CH were selected for further imaging analyses, along with 90 age- and sex-matched subjects without CH. All 180 subjects were verified to have no pre-existing myeloid neoplasm.

#### Image Learning Analysis (Cohort 2)

Digital blood cell images were acquired using a Sysmex DI-60 scanner. Image analysis of neutrophils from CellaVision (software version 7.0.1, build 19) single-cell images was run in Python 3.11.4 with OpenCV 5.0.0. Neutrophil cell identification and segmentation was conducted with a U-Net deep learning architecture with a ResNet34 encoder using pre-trained ImageNet weights through segmentation-models-pytorch 0.5.0.^35,36^ To acclimate the model to our data we manually labelled 311 CellaVision neutrophil images with the LabelStudio brush tool, including 172 in-house images and 139 publicly available images.^37^ This sample of cell images was determined to be sufficient as validation Dice learning curves showed plateauing segmentation accuracy with additional annotated images. The robustness of cell segmentation was enhanced via image augmentations, including colour adjustment, noise and blur, and random geometric transformations to fine-tune a pretrained model with ImageNet encoder weights. Model training was guided by Dice and binary cross-entropy (BCE) loss scores to optimize segmentation performance. When evaluated on a validation set of 31 held-out neutrophil images, the automated cell outlines achieved a 97% mean intersection-over-union (i.e. spatial overlap) with manually drawn cell boundaries. Binary cell masks from the U-Net model were used to isolate the cell and mask out the extracellular image background. Each neutrophil cell image was then segmented into its nuclear and cytoplasmic compartments based on staining intensity through Otsu thresholding.

Once segmentation was completed, a total of 34 human interpretable features were extracted from each cell, spanning image colour (n=18), texture (n=6), and geometry and morphology (Cellular n=6; Nuclear n=4).^38^ Colour features were taken from the isolated cytoplasm mask and were processed and analyzed in the LAB colour space (L, Lightness; A, Red-Green Axis; B, Blue-Yellow Axis), which is highly effective for isolating colour variations in blood smear images.^38^ To ensure consistency across different stains and backgrounds, Otsu thresholding was applied to the background mask, and the average LAB pixel value was subtracted from the cytoplasm mask. Each of the three channels of the normalized LAB colour space were converted to grayscale, and the 6 classical first order statistical features (mean, standard deviation, skewness, kurtosis, energy, and entropy) were calculated for each channel. Gray-level cooccurrence matrix (GLCM) features (contrast, dissimilarity, angular second moment [ASM], energy, entropy, and homogeneity) were taken from the grayscale cytoplasm mask to assess image texture.^39^ Cellular geometry was evaluated via diameter, area, elongation, circularity, convexity, and solidity. Nuclear geometric features included area, convexity, solidity, and the number of nuclear lobes. Nuclear lobes were counted with a skeleton graph of the nucleus, which was used to derive a width function where marked width reductions represented lobular separations.^40^

We then curated single-cell images of 720 relatively hypogranular neutrophils from 29 patients (mean age 72.9 years, M:F ratio 0.55) and 900 non-hypogranular (normal + hypergranular) neutrophils from 26 patients (mean age 60.9 years, M:F ratio 0.60) to train classification models to identify neutrophil granularity from blood slide images. The relative granularity of each neutrophil image used for training was assessed by an expert hematopathologist. Cell images were purposefully selected from a diverse range of samples to cover the full spectrum of neutrophil granularity states, spanning both normal and malignant hematopoiesis. Features from each of these images were extracted as described above and input to support vector machine (SVM), random forest, and LASSO logistic classifier models implemented in scikit-learn 1.9.0. Training and testing was performed using 10-fold nested cross validation. The SVM was the strongest performing model with a mean classification accuracy of 95.1% (Supplemental Figures 7-8). The SVM model was then deployed on 14,403 neutrophil images from the 180 selected patients (mean neutrophils/patient = 80, SD = 16) to quantify the proportion of neutrophils in each sample that displayed a relatively hypogranular phenotype. The proportion of relatively hypogranular neutrophils was then compared across CH status using a beta-binomial regression model adjusted for age, sex, and comorbidities using glmmTMB 1.1.11 in R 4.4.2.

#### DNA Sequencing - Bone Marrow Subcohort (Cohort 3)

Genetic testing for myeloid cancer driver mutations was conducted with an established ion semiconductor sequencing assay, targeting 40 genes recurrently mutated in CH and myeloid neoplasms (Oncomine Myeloid Research Assay, ThermoFisher) as described previously.^41^

#### Flow Cytometry (Cohort 3)

Aspirated bone marrow cells were processed on a Becton Dickinson BD FACSLyric™ System, as part of routine investigations and care. All samples were initially analyzed using a cell count and viability tube that included assessment of cytoplasmic granularity via side scatter-A (SSC-A) ratio, representing the ratio of side scatter measurements of granulocytes versus lymphocytes in each sample (as defined on the cardinal SSC-A by CD45 plot). A subset of samples underwent more extensive analyses that included assessments of granulocyte maturation, assessed by measuring cell surface expression of CD34, HLA-DR, CD117, CD11b, CD13, CD16, CD33, CD35, and CD64 to identify the following populations: promyelocytes (CD117+CD11b-), late promyelocytes/premyelocytes (CD117-CD11b-), myelocytes (CD117-CD11b+CD16-), metamyelocytes (CD11b+CD16+), and neutrophils (including band cells; CD11b+CD16+/CD10+). Where indicated, SSC-A ratio was also calculated for these discrete granulocyte populations for patients with pathogenic/likely pathogenic (P/LP) *TET2* variants versus controls with no identified P/LP variants.

### UK Biobank Mortality Analyses

CH status was ascertained in 451,187 individuals without a history of hematologic cancer as previously described.^42^ Mortality data was collected from death certificate records, with infection-related mortality assigned where the primary cause of death listed was of an infectious etiology. Cox proportional hazard regression was used to assess the association of CH status with all-cause and infection-related mortality. The *MPO* rs2333227 SNP was modeled as an additive term (i.e. 0, 1, or 2 T alleles per person) in interaction analyses. Models were adjusted for age at enrollment, age-squared, sex, smoking history, and 10 principal components of genetic ancestry.

### Statistical Analysis

All analyses were performed in R 4.4.2 and all figures were generated with ggplot2 4.0.2 unless otherwise stated.

## Data Availability Statement

Requests for access to raw or processed study data will be granted upon reasonable request to the corresponding author, Michael J. Rauh. CH calls for UK Biobank participants are available to authorized researchers as of data release version 20 through the UK Biobank Research Analysis Platform.

## Acknowledgements

This work was supported by the Canadian Cancer Society (Research Institute Innovation Grant # 702518, RB and MJR), the Leukemia and Lymphoma Society of Canada (Blood Cancer and AI Catalyst Grant, MJR), the PSI Foundation (PSI Research Trainee Fellowship, EKC), and the Canadian Institutes for Health Research (CIHR Project Grant # 175032, MJR; Vanier Canada Graduate Scholarship, MMB). This research has been conducted using the UK Biobank Resource under Application Numbers 43397 and 101583. We thank Brooke Ring-Snetsinger for assistance with RNA extractions, Eshita Sharma for assistance with RNA sequencing, Erin Allen and Marie Guthrie from the KHSC Core Hematology and Immunology Labs for assistance with blood slide imaging and flow cytometry analyses, and members of the VANTAGE Genomics Core Laboratory at the Vanderbilt University Medical Centre.

## Authorship Contributions

**EKC:** Conceptualization, Methodology, Formal Analysis, Investigation, Writing - Original Draft, Writing – Review and Editing, Visualization. **MMB:** Conceptualization, Methodology, Formal Analysis, Investigation, Writing - Original Draft, Writing – Review and Editing, Visualization. **YZL:** Investigation, Writing – Review and Editing. **AT:** Formal Analysis, Visualization, Writing – Review and Editing. **AD:** Methodology, Writing – Review and Editing. **CV:** Formal Analysis, Visualization, Writing – Review and Editing. **RNA:** Methodology, Writing – Review and Editing. **AJMM:** Methodology, Writing – Review and Editing. **DG**: Resources, Writing – Review and Editing. **RB:** Resources, Funding Acquisition, Writing – Review and Editing. **GRQ:** Resources, Formal Analysis, Writing – Review and Editing. **JB:** Resources, Supervision, Writing – Review and Editing. **AGB:** Resources, Supervision, Writing – Review and Editing. **MJR:** Conceptualization, Methodology, Formal Analysis, Investigation, Resources, Funding Acquisition, Project Administration, Writing – Review and Editing.

