## Supplemental Tables & Figures for "*TET2*-mutant clonal hematopoiesis alters neutrophil maturation and granularity"

**Supplemental Table 1:** Clinical characteristics of participants profiled with whole blood RNA sequencing (Cohort 1)

| Variable | CH (n=21) | No CH (n=9) |
|---|---|---|
| Age, Mean (SD) | 80.9 (7.7) | 78.4 (9.1) |
| Sex, % Male | 38.1% | 33.3% |
| ECOG Score, Median (IQR) | 0 (0, 1) | 0 (0, 0) |
| Charlson Comorbidity Index, Median (IQR) | 1 (0, 3) | 1 (0, 1) |

**Supplemental Table 2:** Differentially Expressed Genes in Whole Blood Samples from Participants with CH versus no CH

| Gene | $Log_2$ Fold Change | P Value | Adjusted P Value |
|---|---|---|---|
| *DEFA4* | 1.91 | 9.5e-08 | 0.003 |
| *ENSG00000294363* | 0.33 | 6.5e-07 | 0.009 |
| *CRISP3* | 1.10 | 6.5e-07 | 0.009 |
| *BPI* | 0.57 | 2.4e-06 | 0.019 |
| *LTF* | 0.49 | 6.8e-06 | 0.042 |
| *GNLY* | -0.17 | 1.9e-05 | 0.062 |
| *ENSG00000303090* | 0.35 | 7.9e-06 | 0.094 |
| *ZIC5* | 0.15 | 3.5e-05 | 0.094 |
| *CXXC4* | 0.13 | 3.6e-05 | 0.094 |
| *FOXF2* | 0.18 | 3.6e-05 | 0.094 |
| *PLBD1-AS2* | 0.12 | 4.4e-05 | 0.094 |
| *MMP8* | 0.16 | 4.7e-05 | 0.094 |
| *SIPA1L1* | 0.14 | 4.9e-05 | 0.094 |
| *ENSG00000286364* | -0.14 | 6.4e-05 | 0.094 |
| *DTHD1* | -0.14 | 3.3e-05 | 0.094 |
| *TRAV24* | -0.23 | 2.4e-05 | 0.094 |
| *TRAV38-2DV8* | -0.16 | 4.4e-05 | 0.095 |
| *DENND11* | 0.13 | 5.9e-05 | 0.096 |
| *ABHD17A* | -0.14 | 6.3e-05 | 0.096 |
| *CTSL* | -0.12 | 6.1e-05 | 0.096 |
| *S1PR5* | -0.11 | 6.6e-05 | 0.099 |

**Supplemental Table 3:** Differentially Expressed Genes in Whole Blood Samples from Participants with *TET2*-mutant CH versus no CH

| Gene | $Log_2$ Fold Change | P Value | Adjusted P Value |
|---|---|---|---|
| *LTF* | 1.88 | 4.8e-08 | 0.001 |
| *DEFA4* | 2.12 | 7.3e-08 | 0.001 |
| *CRISP3* | 0.67 | 2.1e-06 | 0.022 |
| *RAP1GAP* | 0.72 | 3.3e-06 | 0.022 |
| *BPI* | 0.37 | 4.8e-06 | 0.025 |
| *FGFBP2* | -0.38 | 6.7e-06 | 0.025 |
| *ENSG00000302062* | -0.88 | 2.9e-06 | 0.025 |
| *ENSG00000303090* | 0.49 | 2.6e-06 | 0.032 |
| *HLA-V* | 0.30 | 5.8e-06 | 0.032 |
| *ENSG00000306197* | 0.38 | 7.8e-06 | 0.032 |
| *MMP8* | 0.41 | 1.1e-05 | 0.032 |
| *DTHD1* | -0.28 | 9.0e-06 | 0.032 |
| *MXRA7* | -0.25 | 2.2e-05 | 0.057 |
| *ENSG00000301495* | 0.25 | 3.5e-05 | 0.084 |

**Supplemental Table 4:** Differentially Expressed Genes in Whole Blood Samples from Participants with *DNMT3A*-mutant CH versus no CH

| Gene | $Log_2$ Fold Change | P Value | Adjusted P Value |
|---|---|---|---|
| *DENND11* | 0.03 | 4.3e-06 | 0.027 |
| *TSPAN7* | 0.02 | 1.4e-05 | 0.059 |
| *ENSG00000280614* | -0.01 | 3.1e-05 | 0.091 |
| *ENSG00000280800* | -0.01 | 3.1e-05 | 0.095 |

**Supplemental Table 5:** NeuMap Functional Compartment Mappings of Upregulated Genes in *TET2*-mutant CH

| NeuMap Compartment | Gene Overlap | gProfiler P value |
|---|---|---|
| Immature Neutrophils | *LTF*, *DEFA4*, *CRISP3*, *BPI*, *MMP8* | 0.009 |
| Pre-Neutrophil-like Neutrophils | *LTF*, *DEFA4*, *BPI*, *MMP8* | 0.52 |

**Supplemental Table 6:** Clinical characteristics of participants profiled with peripheral blood slide imaging analysis (Cohort 2)

| **Variable** | **CH (n=90)** | **No CH (n=90)** |
|---|---|---|
| Age, Mean (SD) | 75.9 (11.3) | 75.5 (10.3) |
| Sex, % Male | 56.7% | 57.8% |
| No. of Neutrophils in Blood Slide Image, Mean (SD) | 80.2 (15.8) | 79.9 (16.4) |
| Kidney Dysfunction, % | 27.8% | 27.8% |
| Liver Dysfunction, % | 14.4% | 10% |
| Iron Deficiency, % | 12.2% | 11.1% |

**Supplemental Table 7:** Clinical characteristics of participants profiled with genetic and flow cytometric analysis of aspirated bone marrow (Cohort 3)

| **Variable** | ***TET2* Mutation (n=29)** | ***DNMT3A* Mutation (n=7)** | **Other Driver Mutation (n=31)** | **No P/LP Driver* (n=10)** |
|---|---|---|---|---|
| Age, Mean (SD) | 74.9 (9.0) | 75.0 (6.9) | 72.7 (8.1) | 69.7 (14.7) |
| Sex, % Male | 69.0% | 30.0% | 58.1% | 80.0% |
| CCUS, % | 31.0% | 42.9% | 16.1% | 0.0% |
| MDS or MDS/MPN, % | 55.2% | 42.9% | 83.9% | 20.0%** |
| AML, % | 13.8% | 14.2% | 3.4% | 0.0% |
| Non-Dx, % | 0.0% | 0.0% | 0.0% | 80.0% |

*No pathogenic/likely pathogenic (P/LP) SNV or small indel driver identified by NGS panel

**2 cases of myelodysplastic syndrome (MDS) diagnosed morphologically with no identified P/LP driver mutation (i.e. MDS with increased blasts)

**Supplemental Figures**

**Supplemental Figure 1:** Differential gene expression in whole blood from CH and CH-free participants. Select neutrophil and granule related genes are labelled, and coloured points represent nominal P value < 0.05.

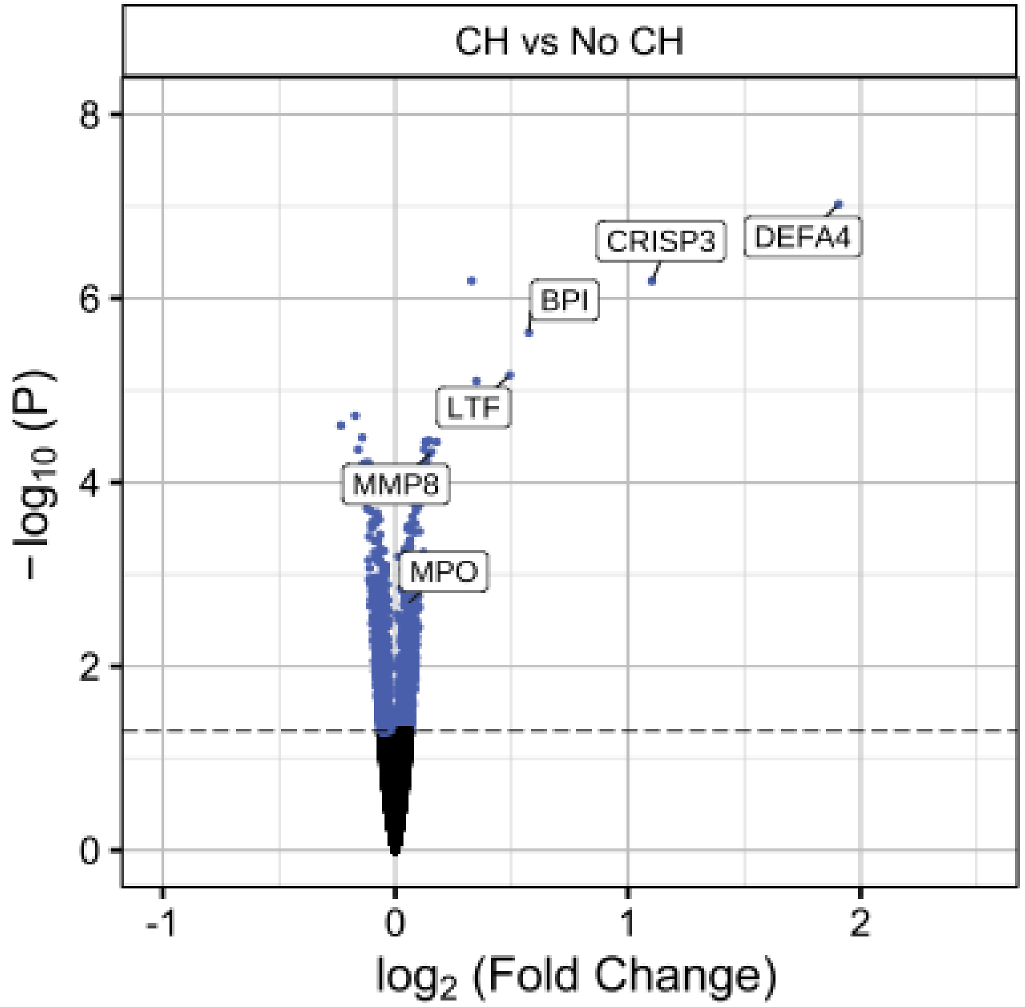

**Supplemental Figure 2:** Differential gene expression in whole blood from *DNMT3A*-mutant CH and CH-free participants. Select neutrophil and granule related genes are labelled, and coloured points represent nominal P value < 0.05.

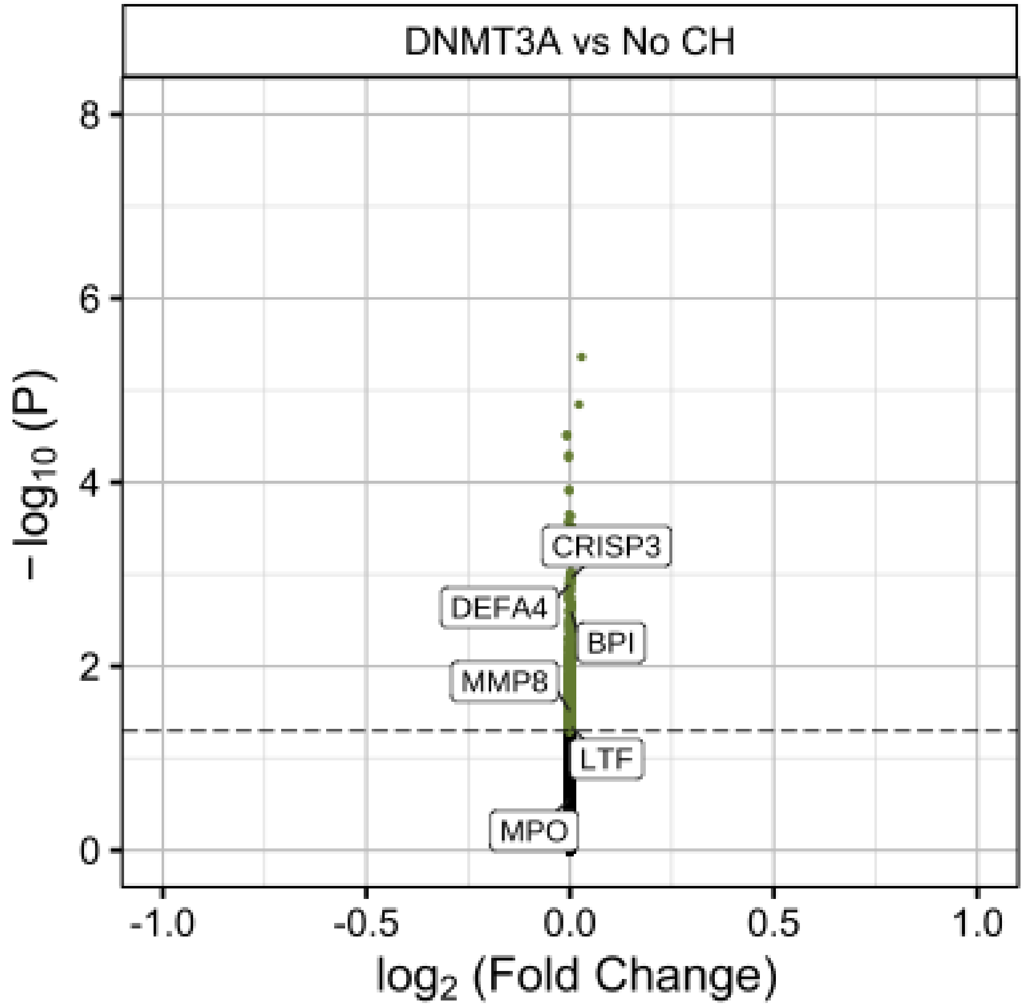

**Supplemental Figure 3:** Absolute neutrophil counts of participants from complete blood count. P values from Wilcoxon rank sum test.

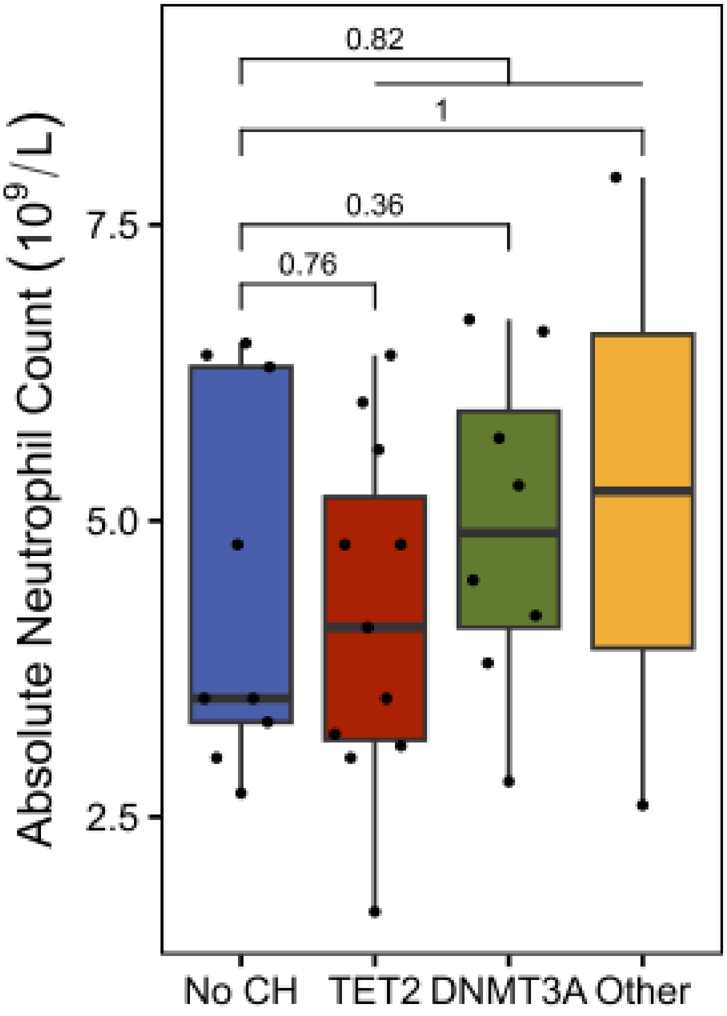

**Supplemental Figure 4:** Transcription factor activity enrichment in *TET2*-mutant CH compared to no CH.

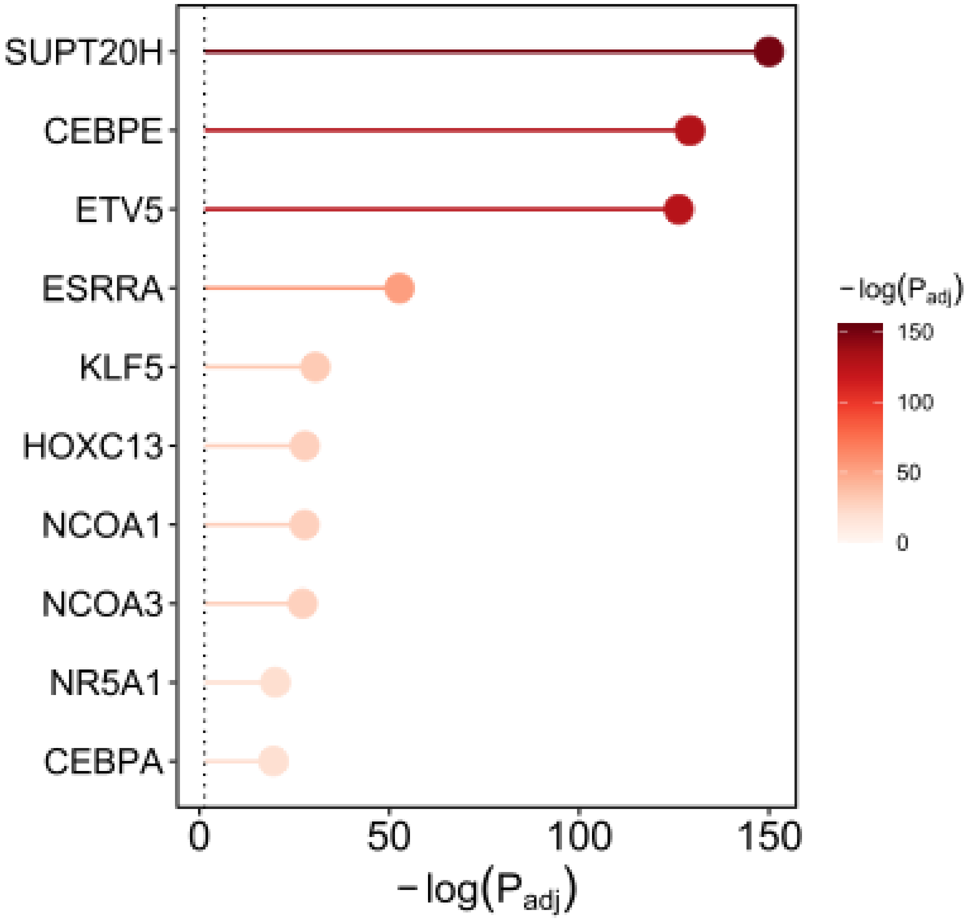

**Supplemental Figure 5:** Principal component plot of image-extracted features of relatively normal/hypergranular and hypogranular blood neutrophils in the model training data.

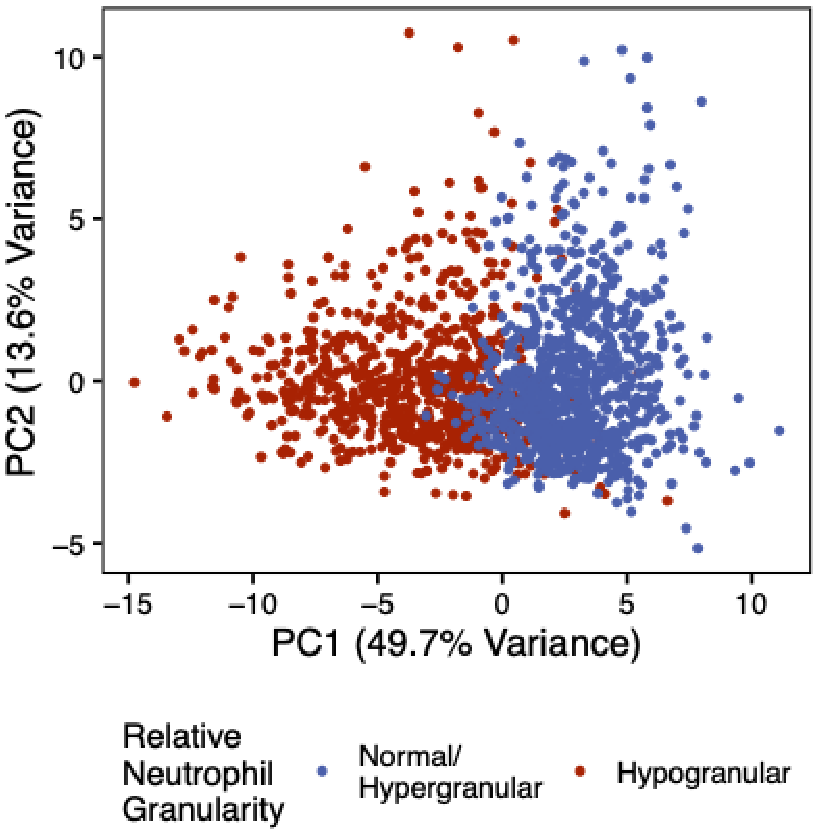

**Supplemental Figure 6:** Mean accuracy (± standard deviation) of normal versus hypogranular neutrophil prediction by the random forest, support vector machine (SVM), and least absolute shrinkage and selection operator (LASSO) logistic classifiers.

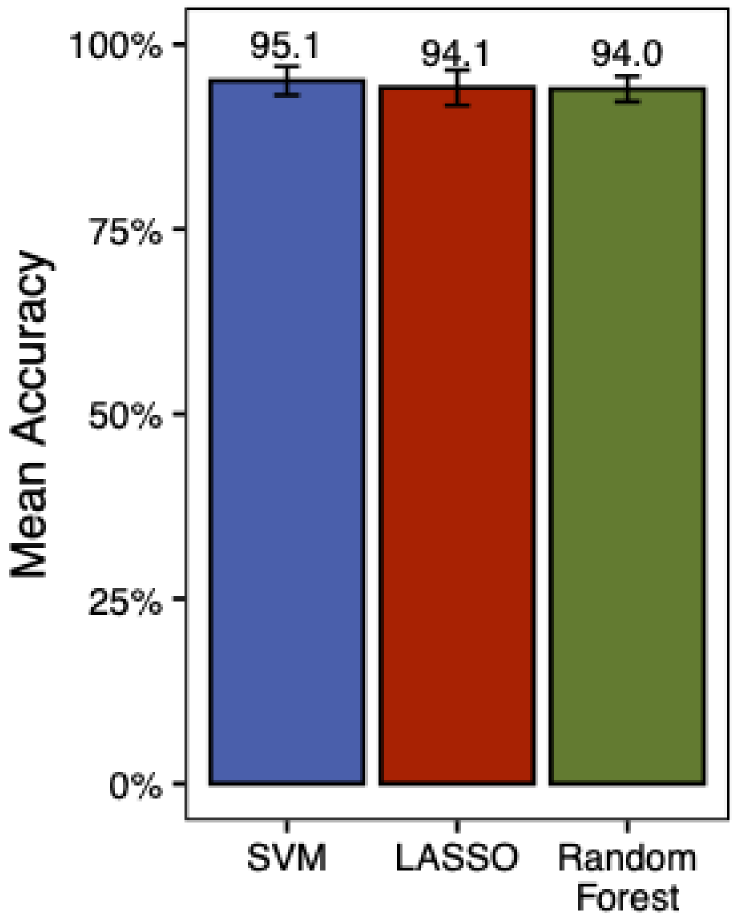

**Supplemental Figure 7:** Image feature weightings used by the support vector machine neutrophil granularity classifier model.

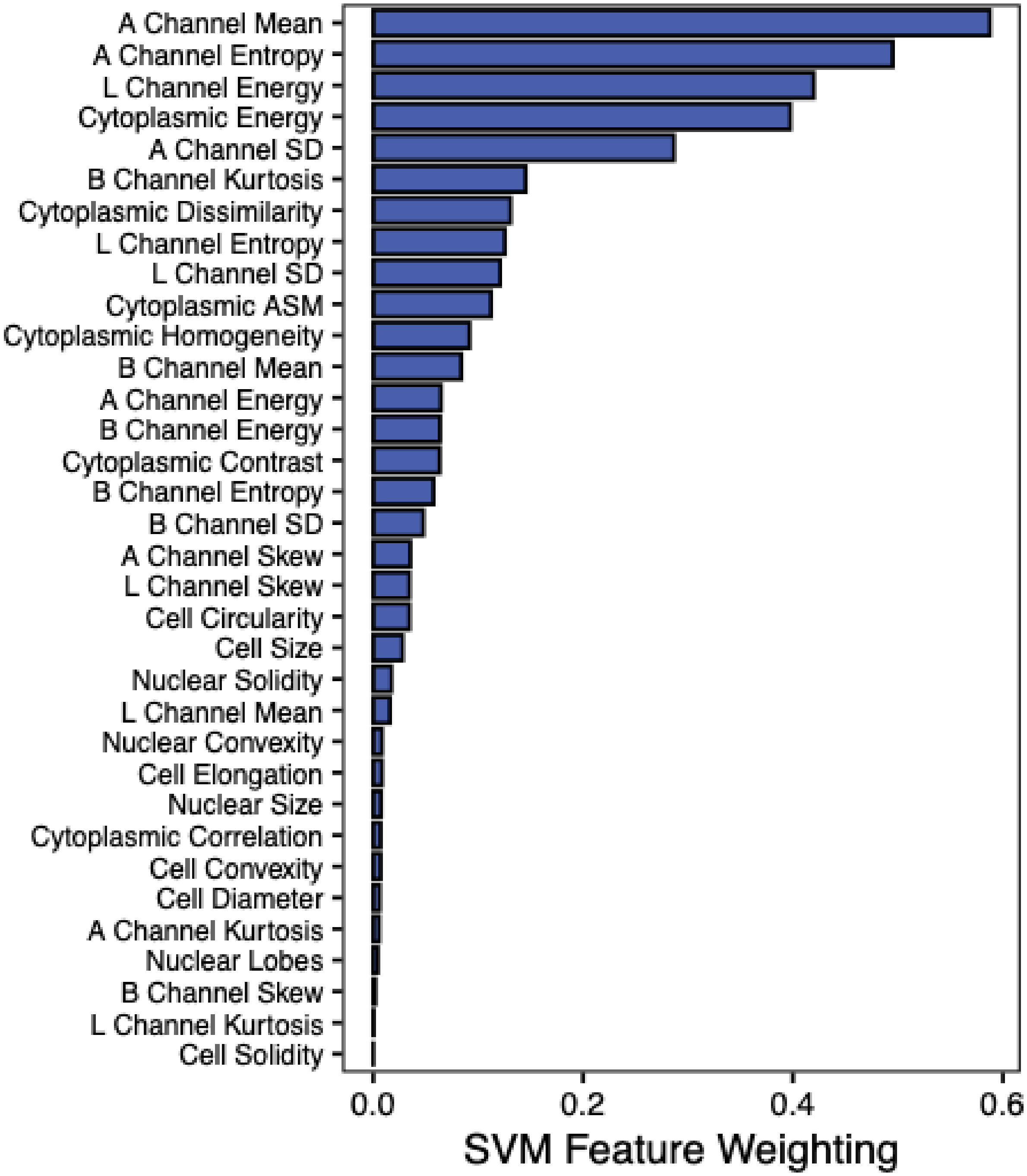

**Supplemental Figure 8:** Correlation of bone marrow side scatter-A (SSC-A) ratio with *TET2* variant allele fraction (VAF) dosage (i.e. cumulative VAF of all *TET2* mutations in the sample). Trendline beta (slope) and intercept, and P value calculated with linear regression adjusted for age and sex.

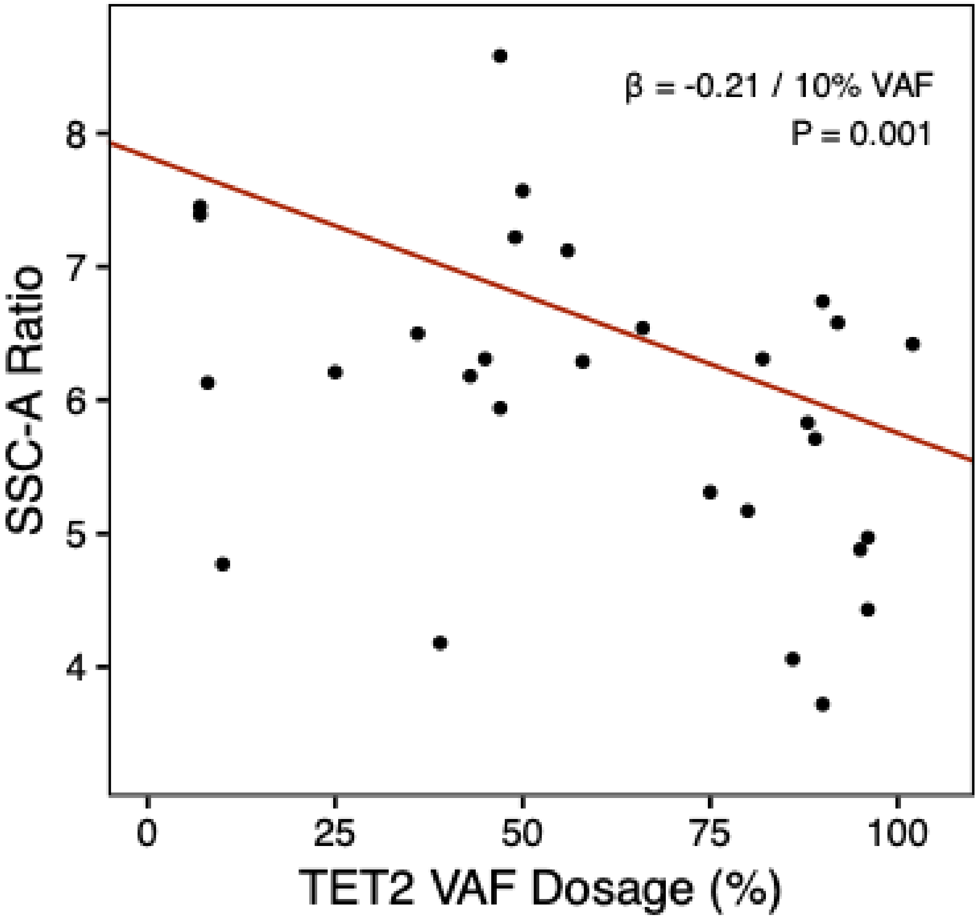
